# Incident hypertension in Urban Slums of Central India: A prospective cohort study

**DOI:** 10.1101/2020.11.30.20240663

**Authors:** Abhijit P Pakhare, Anuja Lahiri, Neelesh Shrivastava, Ankur Joshi, Sagar Khadanga, Rajnish Joshi

**Author notes:** Corresponding Author- Dr Rajnish Joshi, Department of Medicine, All India Institute of Medical Sciences, Bhopal.

## Abstract

**Background:** National Program for prevention and control of cancer, diabetes, cardiovascular diseases and stroke (NPCDCS) in India envisages annual screening of adults over age of 30 years for hypertension. It is followed by confirmation and further linkage for treatment and health promotion interventions. We aimed to estimate incidence rate of hypertension and to identify risk factors for same, so that it is useful for program implementation.

**Methods:** We established a cohort of adults residing in urban slums of Bhopal, who were registered in a baseline cardio-vascular risk assessment survey, which was performed between November-2017 and March-2018. Blood pressure assessment was done at-least thrice at baseline for diagnosis of hypertension, which was defined as SBP >/= 140 mm Hg or DBP >/= 90 mm Hg on two or more occasions. Participants who did not have a diagnosis of hypertension, were followed up during April-June 2019.

**Results:** Of the 5673 participants assessed at baseline, 4185 did not have hypertension of which 3199 (76.4%) were followed up after a median on 1.25 years (IQR 1.08-1.60) and a total of 170(5.31%) individuals were detected with incident hypertension. Overall incidence rate of HTN was 4.1 (95%CI 3.54-4.75) per 100 person-years of follow-up. On multi-variate analysis, age (RR 1.79; 95%CI 1.1-2.92 for age > 60 years) and being illiterate (RR 1.9; (95%CI 1.35-2.69) were significant predictors of incident hypertension. Individuals who had pre-hypertension at baseline also had a significantly increased risk of developing hypertension (RR 3.02; 95%CI 2.06-4.44).

**Conclusions:** We found that incidence of hypertension in urban slums of central India is higher with increasing age and in men. Illiteracy and prehypertension are other determinants. We also demonstrate feasibility of establishing a cohort within the public-health delivery system, driven by efforts of Community Health Workers.

## Introduction

Hypertension is a leading cause of cardiovascular diseases globally and in India. About a third of all urban and a fourth of all rural adults have hypertension.^[1]^ Previous studies by our group have estimated prevalence of hypertension and its risk factors in state-wide representative survey of Madhya Pradesh. We found that prevalence to be 25.7% in urban, and 20.8% in rural areas.^[2]^ These numbers translate into more than 200 million individuals with hypertension in India.^[3]^ Given this huge burden, National Program for Cancer, diabetes, cardiovascular diseases and Stroke (NPCDCS) launched by government of India envisages annual screening for hypertension in all adults above 30 years of age.^[4]^ Once such a screening is fully operationalized, individuals with newly detected hypertension are likely to add-on to the overall burden. There are only a limited studies on incidence of hypertension from India. In a previous cohort of 300 individuals from southern Indian state of Kerala, India, nearly one-fourth individuals developed hypertension in seven years of follow up from 2003-2010.^[5]^ This 3.3% annual incidence, was the only community based benchmark from India, till publication of a multi-city CARRS Study (Chennai, Delhi, and Karachi) in 2017 that provided an annual incidence estimate of 8.2%.^[6]^ A recently published cohort from eastern state of West Bengal has reported incidence of hypertension to be 5.9% for women, and 7.9% for men respectively.^[7]^

Rise in incidence has led to increase in prevalence of hypertension, and bridging of rural-urban gap.^[8]^ It is important to identify risk factors of such incident hypertension beyond age, in case the tide needs to be stemmed. Previous studies have identified elevated blood pressure (previously known as pre-hypertension) as a key risk factor, which simply indicates a transition from a normal to hypertensive state.^[5,6]^ Obesity, smoking, alcohol-use and dysglycemia are other reported significant factors in these two studies from India. Lack of physical activity and increase in alcohol use have been identified as key-risk factors in two recent meta-analyses.^[9,10]^ The evidence base of incident-hypertension and its risk factors is sparse and there is a need to better understand relationships in cohort studies, especially in vulnerable population subgroups.

Two-thirds of population of India is young, and more than half of all adults residing in urban areas are economically disadvantaged.^[11]^ These population subgroups are most vulnerable to develop incident hypertension, contributing to overall cardiovascular disease burden. The current study is designed to estimate incidence of hypertension and its predictors on follow-up. In addition to project future anticipated burden of disease, and identification of vulnerable groups, Incidence information is also important to assess operational adequacy of NPCDCS in the envisaged annual follow up surveys.

## Methods

### Design and Ethics Statement

We established a cohort of adults residing in urban slums of Bhopal to estimate incidence of hypertension. Detailed methods of the study are described previously ^[12]^ Baseline and follow up assessment protocol was approved by the Institutional Human Ethics committee of All India Institute of Medical Sciences, Bhopal. (IHEC-LOP/2017/EF00045) All participants provided a written informed consent prior to baseline assessment.

### Setting

We identified a total of 16 Urban slum clusters in Bhopal, a city located in Central India. Accredited social health workers (ASHAs), one from each cluster was trained to perform cardiovascular disease (CVD) risk assessment, follow-up of the participants, and to improve their linkages with public health system. They were supported by a team of study physicians and supervisors, for hand-holding and confirmation of hypertension status.

### Participants

All adults residing in these areas were invited for participation in a baseline cardio-vascular risk assessment survey, which was performed between November 2017 and March 2018. The primary purpose of the baseline survey was to identify individuals at higher CVD-risk (hypertension, diabetes mellitus) and improve their linkages to primary care facilities. Blood pressure assessment was done at-least thrice at baseline for diagnosis of hypertension, which was defined as SBP >/= 140 mm Hg or DBP >/= 90 mm Hg on two or more occasions.

All the participants of baseline survey, who did not have a diagnosis of hypertension, were invited for follow up. Individuals who were pregnant, or those who refused for a follow-up assessment were excluded. There were no other exclusions. Follow up survey was performed between April and June 2019.

### Procedures

Both baseline and follow up measurement of blood pressure was performed using same methodology. We measured blood-pressure using a digital sphygmomanometer (Omron digital apparatus, Model 7200, Kyoto, Japan) with standard size adult cuff. All measurements were done with participants in a sitting position, with well-supported arm and back. An average of three readings obtained one minute apart was recorded as blood pressure at that time. These measurements were obtained at home by ASHAs. Blood pressure was verified for all individuals who had values for SBP>/= 140 mm Hg or DBP >/= 90 mm Hg, and for 10% of all other individuals by a trained supervisor. All individuals who had elevated blood pressure, had their status verified by a study physician at a primary-care facility, before classifying them as having newly diagnosed hypertension.

### Statistical Analysis

Baseline data was collected on mobile phone based application (CommcareHQ), it was then exported in excel and then cleaned and analysed in R software.^[13]^ Information on baseline variables (age, gender, education, wealth-quintiles, smoking status, alcohol consumption, physical activity level, body-mass index, waist circumference, glycemic status) was abstracted from this dataset. All individuals who did not have a diagnosis of hypertension at baseline, but had their SBP between 120 and 139 or diastolic BP between 80 and 89 were classified as having elevated blood pressure (or pre-hypertension). All individuals who had their random blood sugar values of 140mg/dl or greater at baseline, were classified as having dysglycemia. Incidence rate of hypertension and its confidence interval were estimated by using *binom*^[14]^ package in R software which uses binomial distribution. It is expressed as incidence per 100 person years of follow up. *gtsummary*^[15]^ package in R software was used to create summary table grouped by presence or absence of incident hypertension. Comparison of distribution of socio-demographic and clinical factors among those who developed incident hypertension and those who didn’t was done by using Chi-square or t-test appropriately. Development of incident hypertension in a given time is a binary variable. To identify the independent predictors of developing incident hypertension, we have performed relative risk regression by using maximum likelihood regression for log-binomial models. *logbin*^[16]^ package in R software was used for this purpose. Logbin package provides different algorithms for fitting log-link binomial generalized linear model (relative risk regression), allowing stable maximum likelihood estimation and obeying parameter constraints.^[16]^ We first entered socio-demographic and clinical variables individually to estimate relative risk and confidence interval while adjusting for follow-up time. Then we have selected variables with p<0.20 for entering in to relative risk regression model. We have used “*CEM*” algorithm and “*squarem*” method for acceleration in *logbin* package. Then adjusted relative risk and its confidence interval are presented in results.

## Results

Of the 6174 participants at approached at baseline, two blood pressure readings on separate occasions were available for 5673 participants. Out of these 4185 participants did not have hypertension at baseline and were considered as cohort base. Of these 3199 (76.4%) were followed up after a median on 1.25 years (IQR 1.08-1.60). (Figure 1) Amongst this cohort of 3199 individuals, most were below 45 years of age (2013; 62.9%), were women (1937; 60.5%), had a BMI below 25kg/m2 (1708; 53.4% and were classified as sedentary (2373; 74.2%) based on leisure time physical activity levels.(Table 1) Supplementary Table-1 shows comparison of baseline characteristics among those who were followed and who were lost to follow up. Relatively more men, those belonging to lower wealth quintile, having low BMI and low waist circumference, a non-sedentary lifestyle, tobacco users and alcohol users were more likely to be lost to follow up.

**Table 1:**
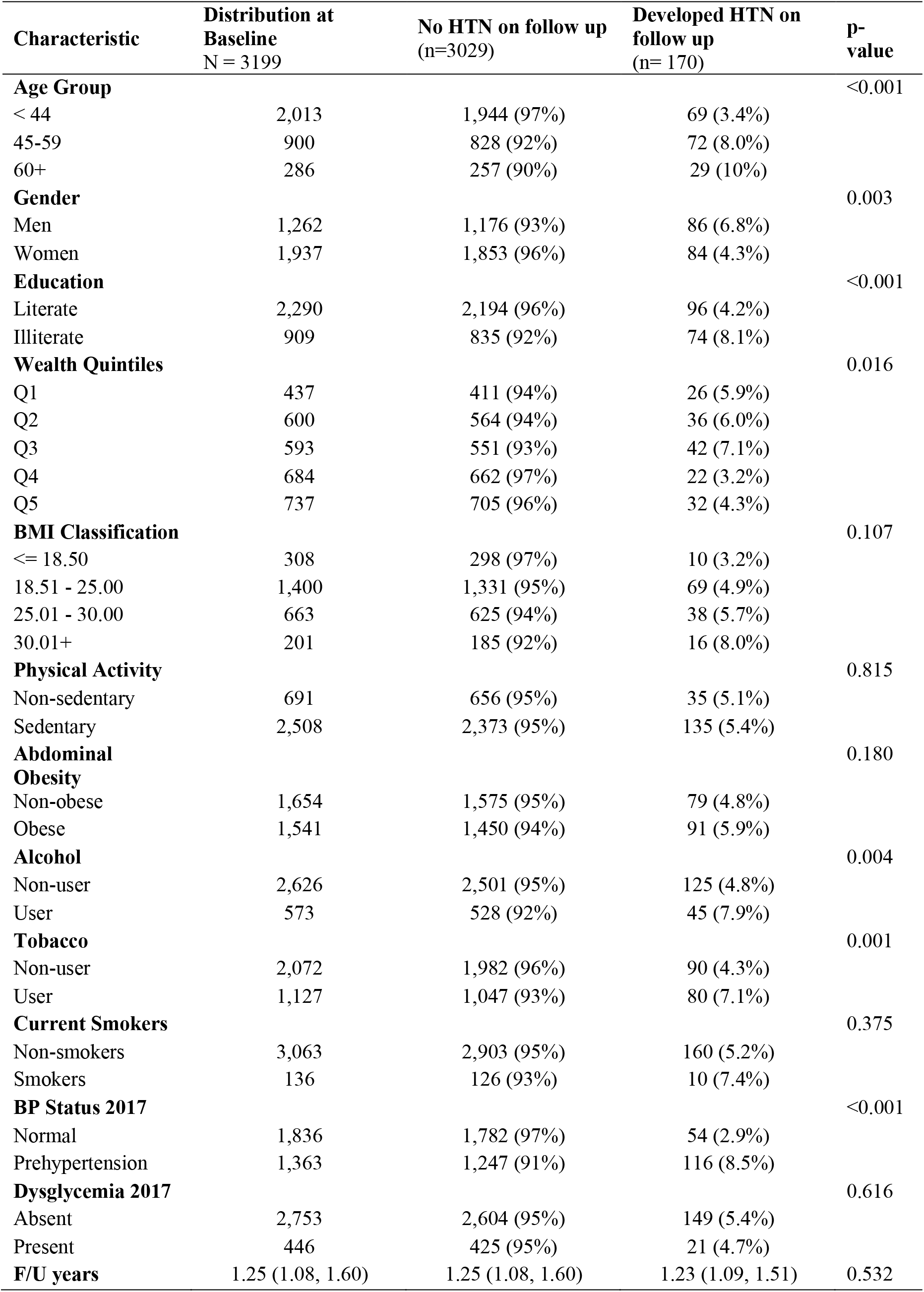
Characteristics of individuals without hypertension at baseline, stratified by hypertension status on follow up (n=3199)

**Figure 1:**
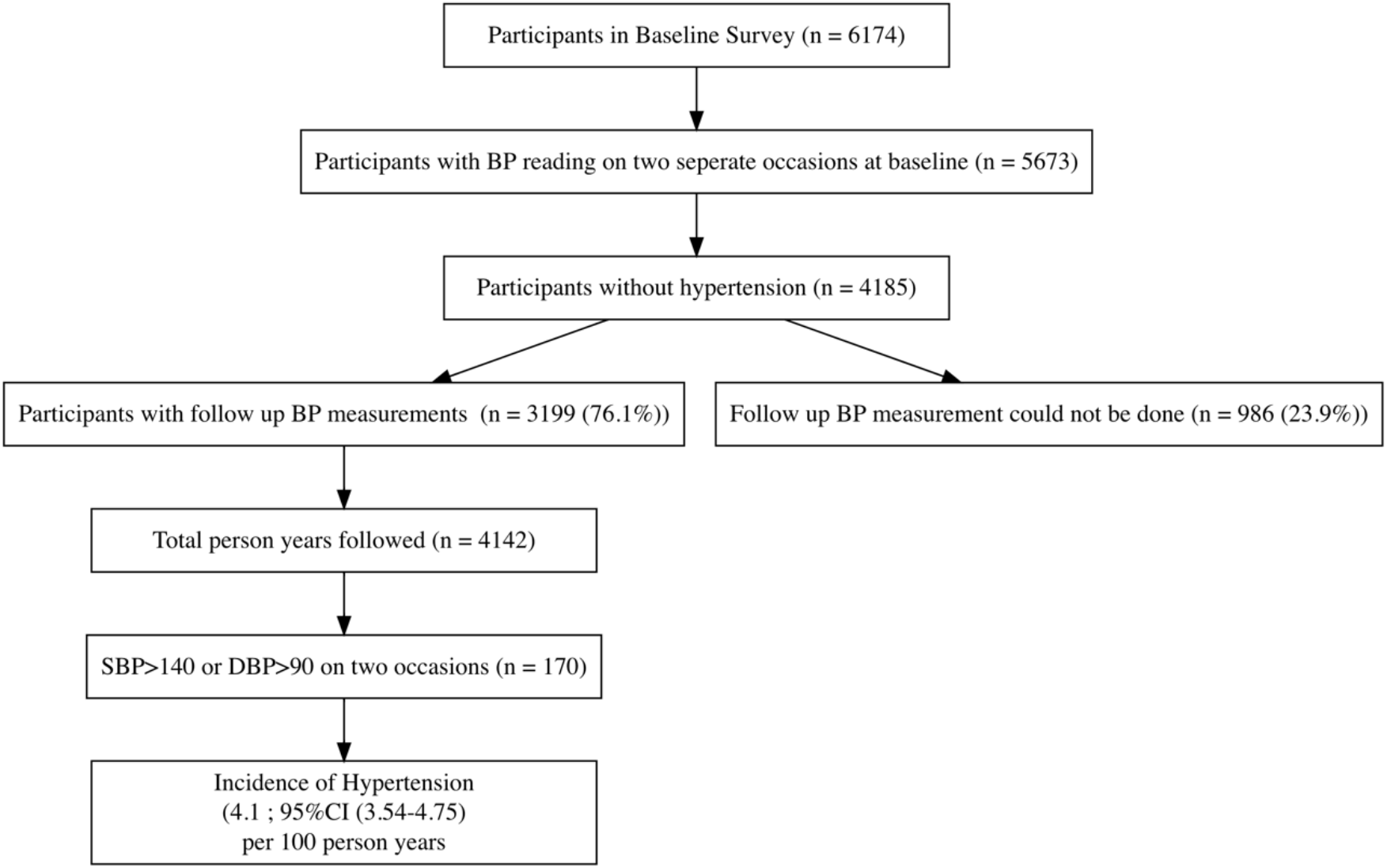
Study Flow.

A total of 170 individuals (5.31%) were detected with a new onset hypertension. Overall incidence of HTN was 4.1 (95% CI 3.54 – 4.75) per 100 person-years of follow up. In all age-groups, incidence was more in men as compared to women. Incidence was highest in men above the age of 60 years (9.36; 95%CI 5.92-14.47) and lowest in women below 45 years of age (2.22; 95%CI 1.63-3.03) per 100 person-years. (Figure 2) SBP values had a clear upward trend with age. SBP values on follow-up were lower than baseline in most age-bands. (Figure 3)

**Figure 2:**
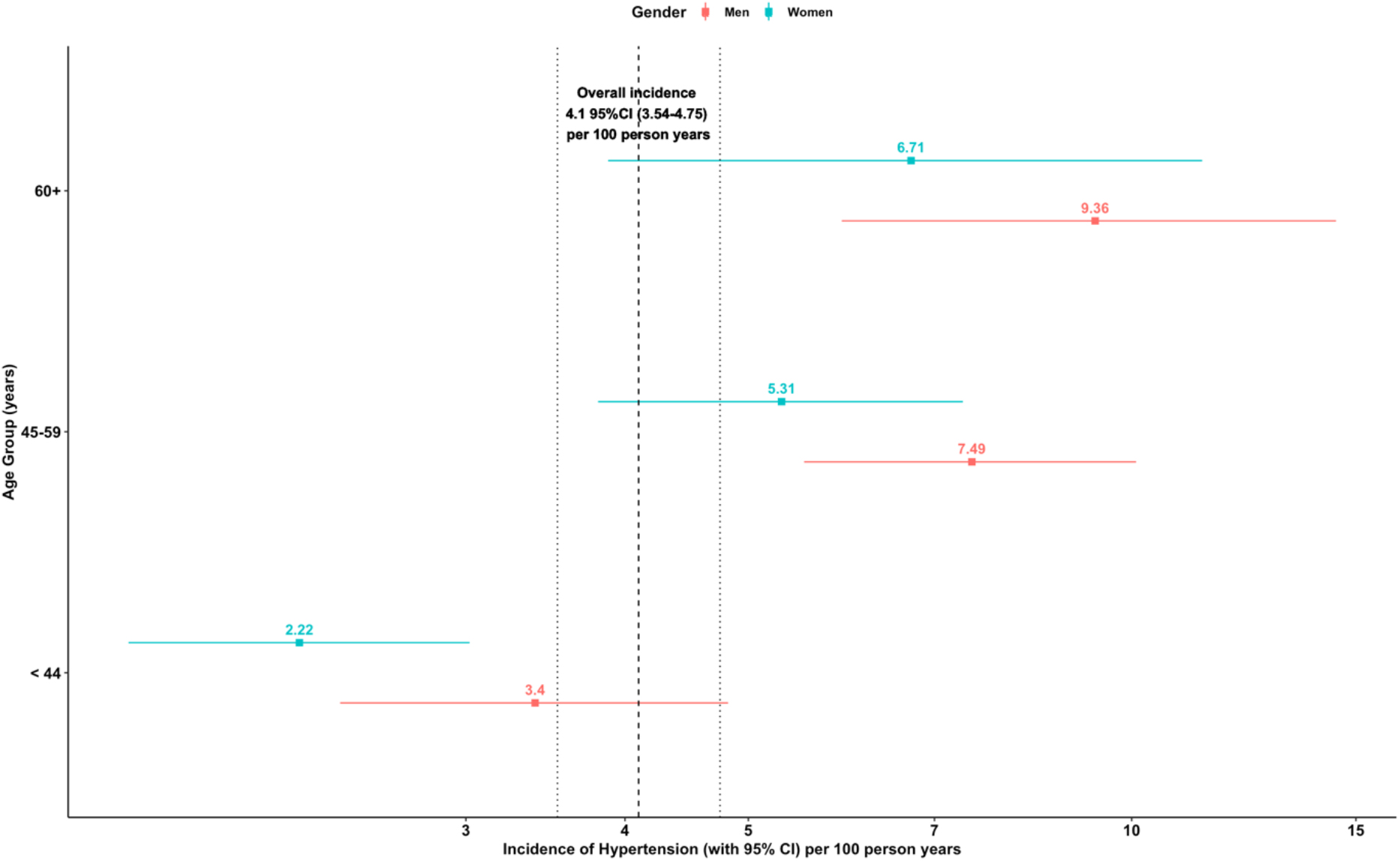
Incidence of Hypertension by age and gender. *Figure depicts incidence of hypertension per 100 person years with its 95% confidence interval of follow up across age group stratified by gender. Central dashed line indicates overall incidence of 4*.*1 per 100 person years while dotted lines on left and right indicates lower and upper limits of 95% confidence interval*.

**Figure 3:**
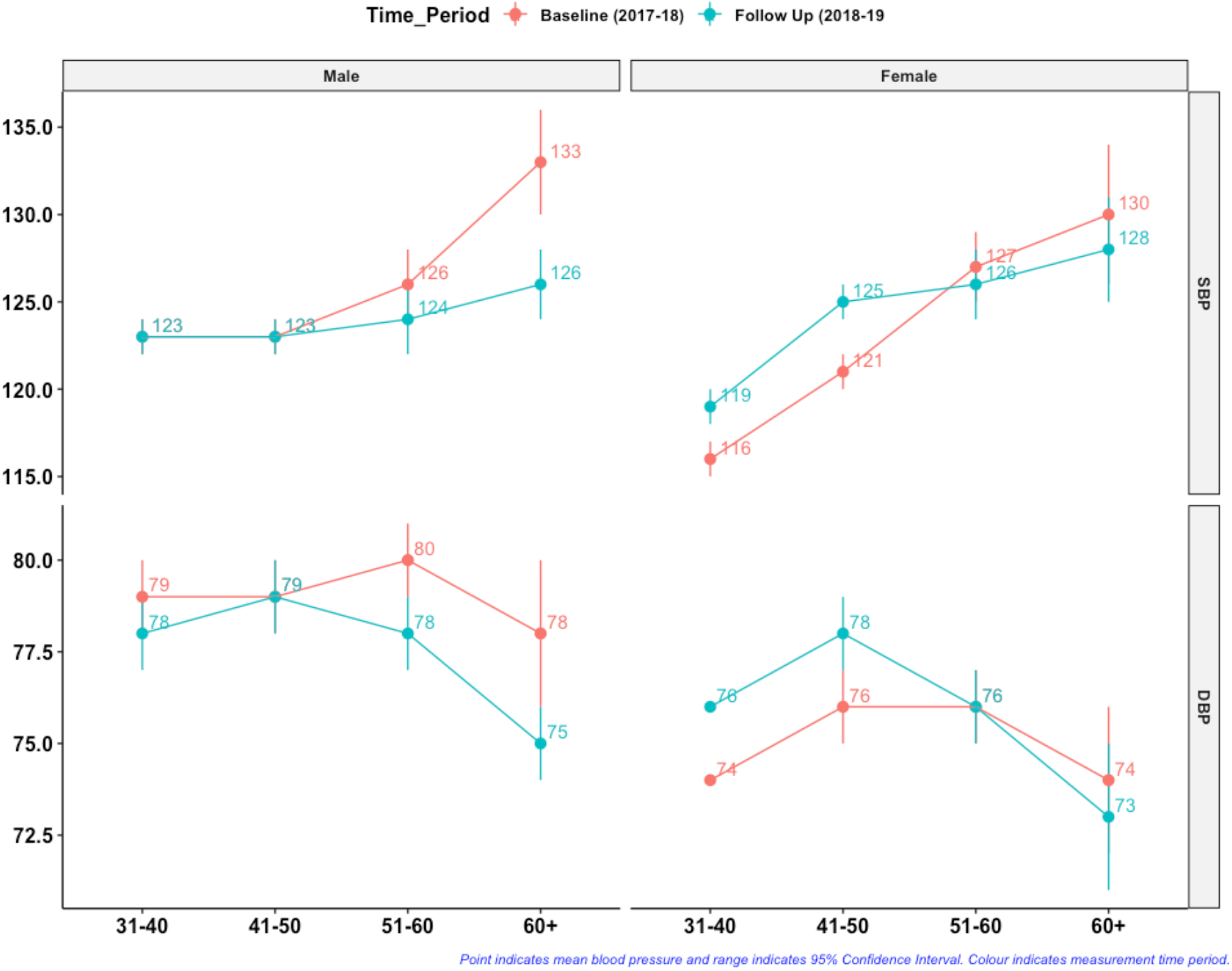
Blood pressure values at baseline and follow up by age-gender categories.

On multi-variate analysis age was a significant predictor of incident hypertension (RR 1.79; 95%CI 1.1-2.92 for age more than 60 years). Amongst modifiable risk factors, BMI greater than 30kg/m2 increased the risk of new onset hypertension (RR 1.93; 95%CI 0.87-4.25), however it was statistically not significant. Individuals who had an elevated blood pressure at baseline (or pre-hypertension) had a significantly increased risk of converting to hypertension on follow up (RR 3.02; 95%CI 2.06-4.44). The risk of new onset hypertension was more in individuals who were not literate (RR 1.9; 95% CI 1.35-2.69), however we did not find a significant risk-gradient with respect to wealth quintiles. (Table 2)

**Table 2:**
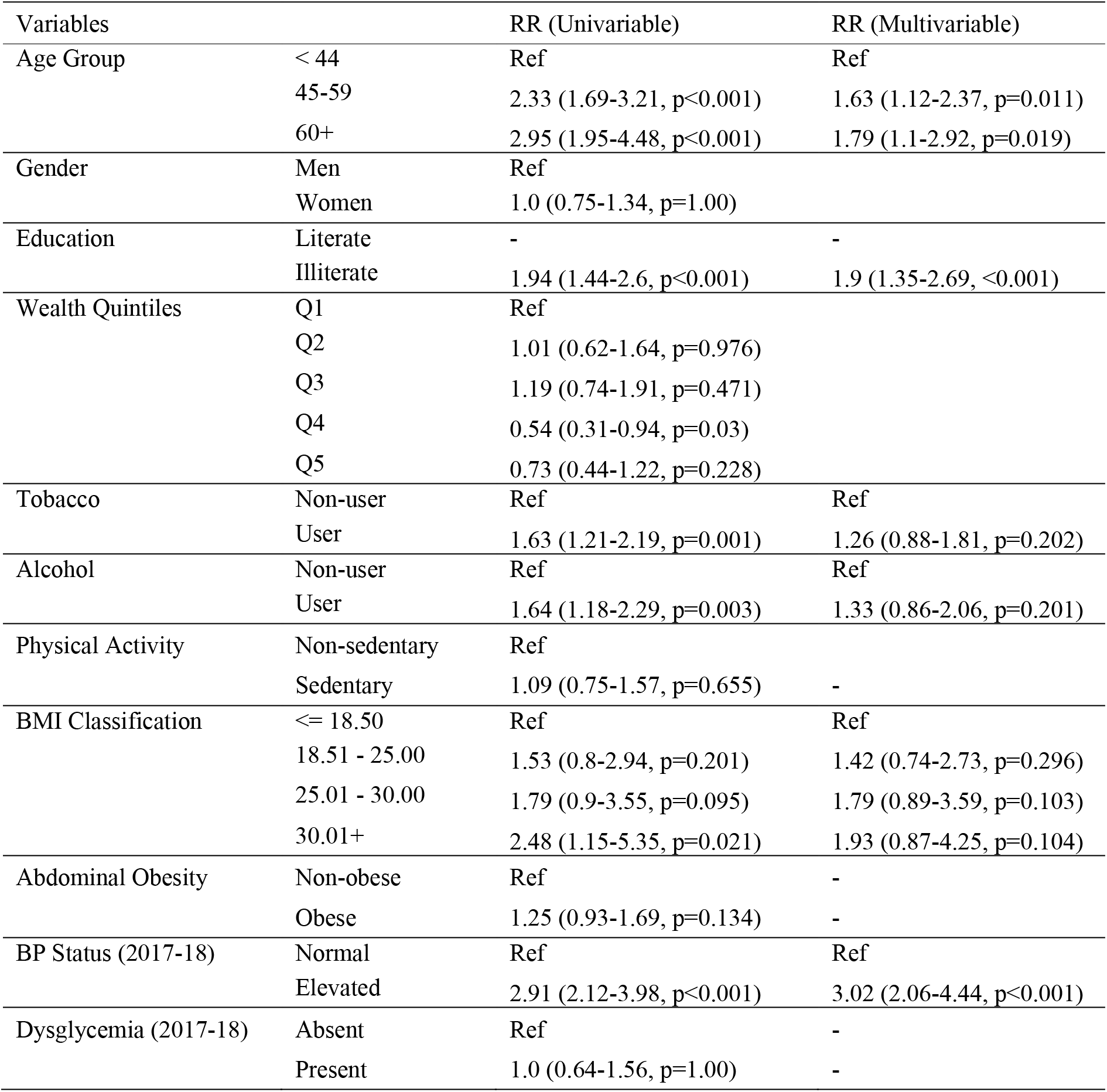
Risk of incident hypertension, Univariate and multivariate relative risk regression analysis (n=3199)

| Variables |  | RR (Univariable) | RR (Multivariable) |
| --- | --- | --- | --- |
| Age Group | < 44 | Ref | Ref |
|  | 45-59 | 2.33 (1.69-3.21, p<0.001) | 1.63 (1.12-2.37, p=0.011) |
|  | 60+ | 2.95 (1.95-4.48, p<0.001) | 1.79 (1.1-2.92, p=0.019) |
| Gender | Men | Ref |  |
|  | Women | 1.0 (0.75-1.34, p=1.00) |  |
| Education | Literate | - | - |
|  | Illiterate | 1.94 (1.44-2.6, p<0.001) | 1.9 (1.35-2.69, <0.001) |
| Wealth Quintiles | Q1 | Ref |  |
|  | Q2 | 1.01 (0.62-1.64, p=0.976) |  |
|  | Q3 | 1.19 (0.74-1.91, p=0.471) |  |
|  | Q4 | 0.54 (0.31-0.94, p=0.03) |  |
|  | Q5 | 0.73 (0.44-1.22, p=0.228) |  |
| Tobacco | Non-user | Ref | Ref |
|  | User | 1.63 (1.21-2.19, p=0.001) | 1.26 (0.88-1.81, p=0.202) |
| Alcohol | Non-user | Ref | Ref |
|  | User | 1.64 (1.18-2.29, p=0.003) | 1.33 (0.86-2.06, p=0.201) |
| Physical Activity | Non-sedentary | Ref |  |
|  | Sedentary | 1.09 (0.75-1.57, p=0.655) | - |
| BMI Classification | <= 18.50 | Ref | Ref |
|  | 18.51 - 25.00 | 1.53 (0.8-2.94, p=0.201) | 1.42 (0.74-2.73, p=0.296) |
|  | 25.01 - 30.00 | 1.79 (0.9-3.55, p=0.095) | 1.79 (0.89-3.59, p=0.103) |
|  | 30.01+ | 2.48 (1.15-5.35, p=0.021) | 1.93 (0.87-4.25, p=0.104) |
| Abdominal Obesity | Non-obese | Ref | - |
|  | Obese | 1.25 (0.93-1.69, p=0.134) | - |
| BP Status (2017-18) | Normal | Ref | Ref |
|  | Elevated | 2.91 (2.12-3.98, p<0.001) | 3.02 (2.06-4.44, p<0.001) |
| Dysglycemia (2017-18) | Absent | Ref | - |
|  | Present | 1.0 (0.64-1.56, p=1.00) | - |

## Discussion

We found that incidence of hypertension in urban slums of central India is progressively higher with increasing age, and more in men as compared to women in each age band. Overall about four new hypertensives are likely to be detected for every 100 individuals followed up in a community for a year. This number is 10 and 8 respectively for men and women above the age of 60 years, but about 3 and 2 respectively for those below 45 years of age. This information will be useful operational indicator for annual follow-up assessments planed in NPCDCS. In addition to age, we found lower education level, and elevated blood pressures at baseline as significant predictors of incident hypertension. These features help us to identify vulnerable groups within the urban slum population, who are more at risk. Most concerning of these are individuals who had an elevated blood pressure (also known as pre-hypertension) at baseline, who are at three times greater risk of converting to new onset hypertension. This group is also numerically largest, constituting about two-fifths of the entire cohort at baseline. While various studies have reported prevalence of hypertension in India, there are only a limited studies on its incidence. In a study conducted in 2010 from Kerala (a high NCD burden state in India), the reported incidence of hypertension was 3.3%.^[5]^ More recently CARRS study from metropolitan cities of Delhi, Chennai and Karachi reported a much higher incidence of about 8.2%.^[6]^ These variations could be a reflection of time-period of the study, its setting, or duration of follow up. Various pre-2010 studies from China and Korea have reported lower incidence rates of 2.3%, 5.2%, and 5.3% respectively.^[17–19]^ However in the same time-period, incidence rates in North-America and Europe were comparatively higher, ranging from 6 to 9% in various age-gender subgroups.^[20–22]^ More recent assessments in the previous decade (Table 3) have suggested lower annual incident hypertension rates in studies from communities in Switzerland^[23]^ and Germany^[24]^ (between 2% and 3%), but higher levels (between 8% and 12%) in other studies from China,^[25]^ and Germany. ^[26]^ Such a heterogeneity indicates dynamicity in incidence rates, indicative of epidemiological transition from lower to higher hypertension prevalence. When higher proportion of individuals are classified as having hypertension, pool of at-risk non-hypertensive individuals shrinks, accounting for lower incidence. For instance, in a study from Germany, the annual incidence was between 8-9% in the first four years of follow up, but reduced to 5-6% in the subsequent five years.^[26]^ It has been observed that incidence-rates are lower with longer follow up periods. With that perspective, of the two previous studies from India, a study from Kerala with 3.3% incidence was a seven year follow up from 2003-10, and CARRS study with 8.2% incidence is a two year follow up from 2015-17. Our study from urban slums of a moderate sized city in India has an intermediate overall incidence rate of 4.45% in a median follow up duration of 1.25 years.

**Table 3:**
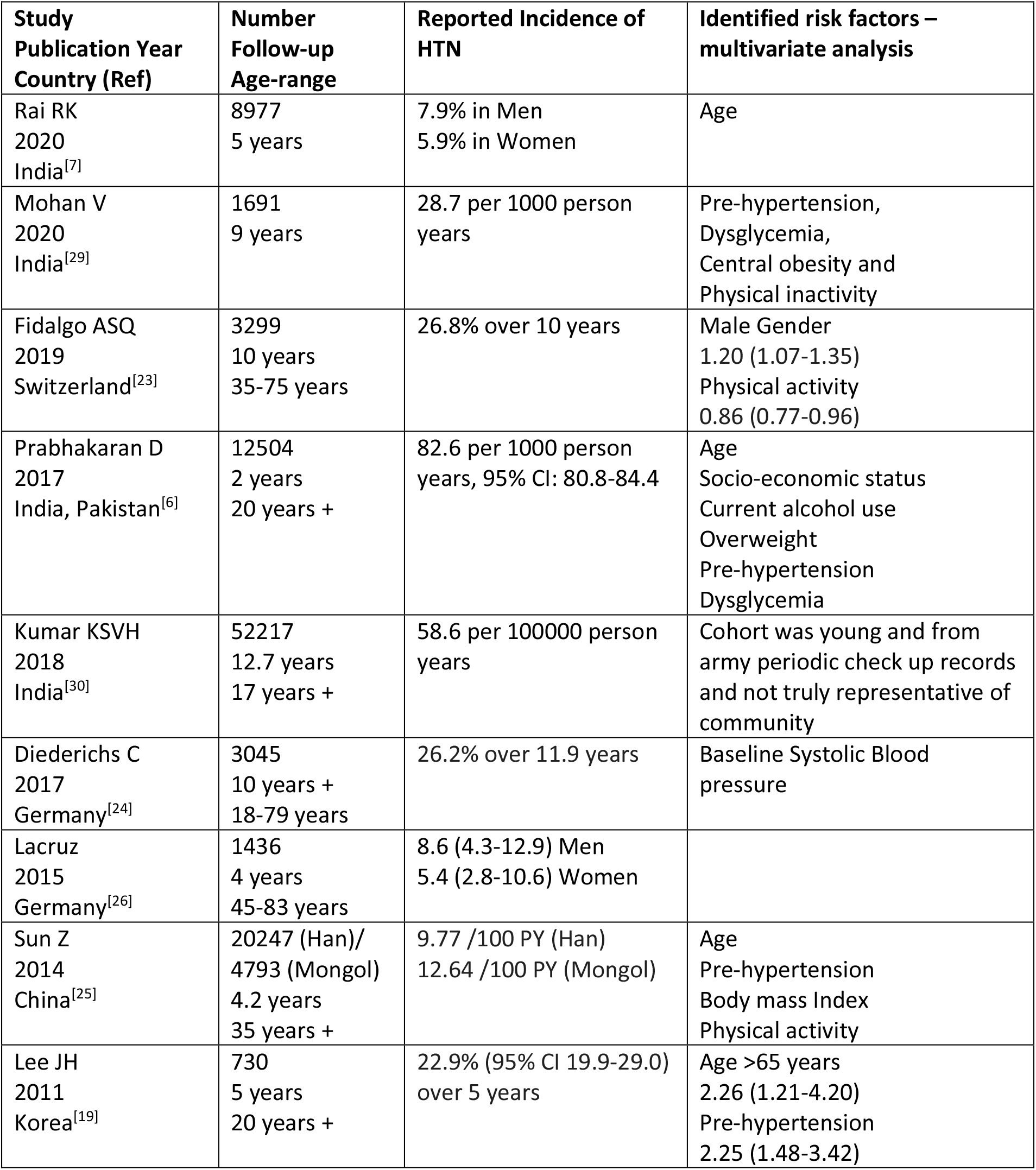
Estimates and determinants of incident hypertension in population based studies published in last decade.

With change in definitions of hypertension, BP below 120/80mm Hg is defined as normal. SBP between 120 and 139, and DBP between 80 and 89 that was previously defined as pre-hypertension is now labelled as “elevated”. While newer hypertension definitions classify blood pressure measured at home of greater than 135/85 as hypertension, European definitions consider this to be equivalent to office blood pressure of 140/90 mm Hg.^[27,28]^ In our study, home blood pressures were measured by ASHAs and office blood pressures were obtained by physicians for confirmation. We have used cut-off of more than 140/90 to define hypertension, and less than 120/80 mm Hg to define normal. Those with intermediary values at baseline are classified as having elevated blood pressure. This intermediary group is prone to a differential misclassification when multiple readings are obtained in individuals. For instance, of the blood pressure values obtained at different occasions, if even one of the three values is above normal (120/80) the individual is classified as having elevated blood pressure. In contrast, we need at-least two values above 140/90 to classify an individual as having hypertension. Thus pool of individuals with “elevated blood pressure” is large, and classification systems force some normotensive and some true-hypertensive individuals in this category. Of all individuals with “elevated blood-pressure” at baseline, 9.8% developed hypertension, constituting more than two-thirds of all newly detected hypertensives. Biological plausibility of this subgroup of individuals developing incident hypertension is well known, and reaffirms application of preventive strategies in them.^[6,19,24]^

Illiteracy was another significant predictor of incident hypertension in our study. Education is a proxy of socio-economic status, and this is an important discriminating variable in our study in urban slum dwellers, where gradient of asset ownership (wealth quintiles) is rather narrow. Smoking and alcohol consumption were also significant on univariate analysis, in our study, but these two are likely to be outcomes of illiteracy and hence intermediary in pathway to causation of hypertension. Previous studies have identified alcohol use as significant risk factor,^[6]^ and a meta-analysis of 20 cohort studies also found that any level of alcohol consumption is associated with increased risk of hypertension (RR 1.51 (95%CI 1.30-1.76) for 3-4 drinks per day, and 1.74 (95%CI 1.35-2.24) for 5 or more drinks per day in men, and 1.42 (95%CI 1.22-1.66) for 3 or more drinks in women).^[9]^ Obesity an indicator of reduced physical activity, was significant predictor on univariate analysis in our study. These two variables have been reported to increase risk of hypertension in recent studies.^[6,23]^ A recent meta-analysis of 22 cohort studies has estimated that every 10 additional metabolic equivalents of leisure time physical activity, reduced risk of incident hypertension by about 6% (RR 0.94 (95%CI 0.92-0.96)).^[10]^

Our results from a vulnerable lower income population adds to the evidence base of determinants of incident hypertension from India. We also demonstrate feasibility of establishing a cohort within the public-health delivery system, driven by efforts of ASHAs who are community health workers in urban slum communities. Blood pressures at baseline as well as on follow up were measured by ASHAs, and a comparable incidence to other similar studies validates their measurement skills. Limitation of the study is a shorter period of follow up, however we have shown that blood pressure change is dynamic, and conversion to a hypertensive state is demonstrable within this interval. Urban slum population is prone to outward-migration, as shown in attrition in cohort numbers, however our overall confidence intervals indicate a reasonable certainty in our point estimates. Results of our study are generalizable to other similar urban slum populations, who share similar built-environments, socio-economic priorities, and vulnerability in terms of access to health-care. We are limited by not being able to generalize our results to other population groups, that do-not share these attributes.

We believe that ours and other similar studies will be able to strengthen implementation of NPCDCS, and help evaluate preventive strategies in shorter time-frames. Since diagnosis of hypertension requires life-long care, our efforts in improving preventive-care, can be measured in terms of annual incidence of hypertension with each passing year. Such estimates are now feasible within programmatic settings, with implementation of annual blood pressure screening in all adults.

## Supporting information

STROBE Statement

## Data Availability

Deidentified raw data will be available on reasonable request to the corresponding author

## Compliance with Ethical Standards

### Ethical approval

**-** The study design was approved by the Institutional Human Ethics Committee of All Inndia Institute of Medical Sciences, Bhopal(Ref: IHEC-LOP/2017/EF00045)

### Informed consent

-Participant Information Sheet in Hindi language was provided to each participant. All participants provided written informed consent prior to initiation of any study procedures.

---

**Supplementary Table-1.**
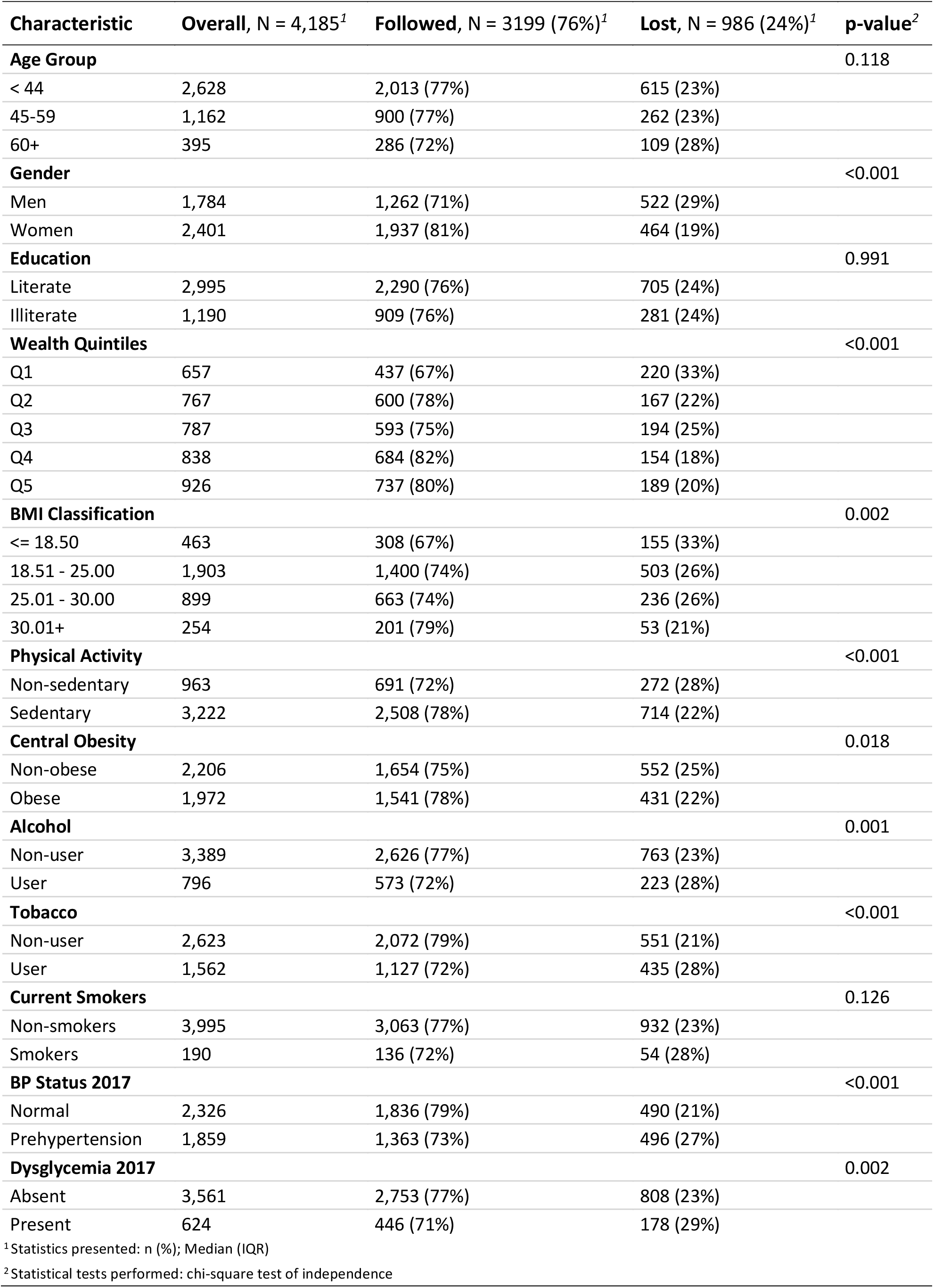

| **Characteristic** | **Overall, N = 4,185<sup>1</sup>** | **Followed, N = 3199 (76%)<sup>1</sup>** | **Lost, N = 986 (24%)<sup>1</sup>** | **p-value<sup>2</sup>** |
| --- | --- | --- | --- | --- |
| **Age Group** |  |  |  | 0.118 |
| < 44 | 2,628 | 2,013 (77%) | 615 (23%) |  |
| 45-59 | 1,162 | 900 (77%) | 262 (23%) |  |
| 60+ | 395 | 286 (72%) | 109 (28%) |  |
| **Gender** |  |  |  | <0.001 |
| Men | 1,784 | 1,262 (71%) | 522 (29%) |  |
| Women | 2,401 | 1,937 (81%) | 464 (19%) |  |
| **Education** |  |  |  | 0.991 |
| Literate | 2,995 | 2,290 (76%) | 705 (24%) |  |
| Illiterate | 1,190 | 909 (76%) | 281 (24%) |  |
| **Wealth Quintiles** |  |  |  | <0.001 |
| Q1 | 657 | 437 (67%) | 220 (33%) |  |
| Q2 | 767 | 600 (78%) | 167 (22%) |  |
| Q3 | 787 | 593 (75%) | 194 (25%) |  |
| Q4 | 838 | 684 (82%) | 154 (18%) |  |
| Q5 | 926 | 737 (80%) | 189 (20%) |  |
| **BMI Classification** |  |  |  | 0.002 |
| <= 18.50 | 463 | 308 (67%) | 155 (33%) |  |
| 18.51 - 25.00 | 1,903 | 1,400 (74%) | 503 (26%) |  |
| 25.01 - 30.00 | 899 | 663 (74%) | 236 (26%) |  |
| 30.01+ | 254 | 201 (79%) | 53 (21%) |  |
| **Physical Activity** |  |  |  | <0.001 |
| Non-sedentary | 963 | 691 (72%) | 272 (28%) |  |
| Sedentary | 3,222 | 2,508 (78%) | 714 (22%) |  |
| **Central Obesity** |  |  |  | 0.018 |
| Non-obese | 2,206 | 1,654 (75%) | 552 (25%) |  |
| Obese | 1,972 | 1,541 (78%) | 431 (22%) |  |
| **Alcohol** |  |  |  | 0.001 |
| Non-user | 3,389 | 2,626 (77%) | 763 (23%) |  |
| User | 796 | 573 (72%) | 223 (28%) |  |
| **Tobacco** |  |  |  | <0.001 |
| Non-user | 2,623 | 2,072 (79%) | 551 (21%) |  |
| User | 1,562 | 1,127 (72%) | 435 (28%) |  |
| **Current Smokers** |  |  |  | 0.126 |
| Non-smokers | 3,995 | 3,063 (77%) | 932 (23%) |  |
| Smokers | 190 | 136 (72%) | 54 (28%) |  |
| **BP Status 2017** |  |  |  | <0.001 |
| Normal | 2,326 | 1,836 (79%) | 490 (21%) |  |
| Prehypertension | 1,859 | 1,363 (73%) | 496 (27%) |  |
| **Dysglycemia 2017** |  |  |  | 0.002 |
| Absent | 3,561 | 2,753 (77%) | 808 (23%) |  |
| Present | 624 | 446 (71%) | 178 (29%) |  |

<sup>1</sup>Statistics presented: n (%); Median (IQR)
<sup>2</sup>Statistical tests performed: chi-square test of independence

